# Developing and piloting a discrete choice experiment to assess stakeholder preferences for community-based exercise programmes after stroke

**DOI:** 10.64898/2026.10.03.26364629

**Authors:** James Smith, Philip Nagy, Carol Holland, Hannah Jarvis, Luis Filipe

## Abstract

**Background:** Community-based exercise may support physical and cognitive recovery after stroke, but the preferences of stakeholders involved in receiving, supporting and delivering these programmes are poorly understood. This study piloted a discrete choice experiment (DCE) that aimed to evaluate stroke survivors’, carers’, clinicians’, and fitness instructors’ preferences for community-based exercise programmes for stroke survivors. The pilot aimed to assess the feasibility of conducting and analysing data for a larger-scale study and inform the sample size for a definitive DCE.

**Methods:** A DCE design was developed using literature reviews, stakeholder consultations, and evaluations of current services. Six attributes were included in the design: location, price, session frequency, individualisation, carer involvement, and recovery. Sixteen Bayesian D-optimal choice sets were developed, then blocked into two sets of eight to reduce cognitive burden. Main-effects were analysed using a mixed logit model, and stakeholder differences were explored using a conditional logit interaction model. Willingness to pay was also calculated.

**Results:** Overall, 161 respondents started the survey and 106 (66%) fully completed it. Stakeholders significantly preferred programmes that were delivered at a leisure centre, lower priced, individualised, had carer-specific sessions and offered greater chances of recovery. The largest willingness-to-pay estimates were £49.41/ month for 80% chance of meaningful recovery vs 20%, and £18.61/ month for leisure centre delivery vs online. Significant preference heterogeneity was identified in the main-effects model. Exploratory stakeholder interactions suggested clinicians were more price sensitive and placed greater importance on recovery and carer-specific sessions than stroke survivors, while fitness instructors placed greater importance on programme location and recovery. Carer interaction estimates were unstable due to the small subgroup sample.

**Conclusions:** The pilot supported the feasibility of the DCE design and analytical approach but identified limitations in the recruitment methods and survey completion. Addition of face-to-face recruitment methods are recommended for a larger-scale study. A larger sample size is required to investigate differences in preferences between stakeholder groups.

## Introduction

Stroke is one of the leading causes of disability worldwide (Feigin *et al*., 2024), leaving survivors requiring long-term support for a range of physical and psychological impairments. The absolute incidence of stroke has been growing since 1990 (Feigin *et al*., 2024), and in the United Kingdom (UK) the number of stroke survivors is forecast to grow to 2.1 million by 2035, which is estimated to cost over £29 billion in associated health and social care costs (King *et al*., 2020). There is evidence that current models of stroke rehabilitation are already struggling to meet the growing demand on services, with only 13.3% of stroke survivors in the UK receiving the recommended amount of rehabilitation (Stroke Association, 2023). The National Health Service has acknowledged these problems in the 2019 long term plan and aims to implement high-intensity rehabilitation interventions that are expected to reduce long term healthcare costs (Alderwick and Dixon, 2019).

Exercise is a low-cost intervention that has demonstrated clinical efficacy for both physical and cognitive rehabilitation after stroke (Saunders *et al*., 2025b, 2025a; Oberlin *et al*., 2017; Kramer *et al*., 2026), and is now recommended by clinical guidelines (Sentinel Stroke National Audit Programme, 2023; NICE, 2023). Exercise can be delivered effectively in a group-format, which can increase capacity of healthcare services, and provide additional psychosocial benefits to stroke survivors (Smith *et al*., 2026b). Exercise services are available to stroke survivors in the UK; however, significant variation in the accessibility, pricing, and delivery of these services has been identified (Smith *et al*., 2026a). There is a need to standardise the provision of exercise services to stroke survivors across the UK, to ensure equal access and opportunity to achieve rehabilitation potential for all stroke survivors, regardless of where they live or level of disability.

To ensure that exercise services are both clinically effective and acceptable, the preferences of stakeholders involved in the programme need to be understood. As stroke survivors are the direct recipients of the interventions, exercise programmes need to be developed based on their needs. However, financial limitations may prevent all these needs being met and understanding what service users are willing to trade-off are essential to balancing acceptability and feasibility. Previous research has highlighted the importance of carers for facilitating exercise in the community and identified gaps in the education and support received during standard rehabilitation (Smith *et al*., 2026b, 2026c). Evidence from the dementia literature suggests that caregiver interventions can improve outcomes for both carers and patients (Cheng *et al*., 2022). Identifying carers preferences will inform their role within community exercise programmes, and ensure appropriate support is provided. Clinicians and fitness instructors are anticipated to deliver the intervention, and ensuring that exercise interventions are aligned to their preferences will help improve intervention fidelity (Johnson *et al*., 2014).

A discrete choice experiment (DCE) is a research method that is used for preference elicitation and quantifying trade-offs (Lancsar and Louviere, 2008). The methodology involves creating hypothetical choice tasks, involving several attributes of interest, that respondents are required to choose from, thereby elucidating their preferences. Previous DCEs have been conducted in the stroke literature (Jolliffe *et al*., 2024), with one specifically aiming to identify stroke survivors preference towards exercise (Geidl *et al*., 2018). Geidl *et al*. (2018) found that stroke survivors had no preference toward exercising individually or in a group; neither at home or in the community. This contrasts with findings from Smith *et al*. (2026c), which found stroke survivors preferred group exercise in the community. The differences may be explained by cultural differences between German and UK stroke survivors, differences in preferences between acute and chronic stroke survivors, or Geidl *et al*. (2018) being underpowered to detect small preferences. There is a need for a well-designed, fully-powered DCE to investigate the preferences of all stakeholders involved with exercise programmes for stroke survivors to aid the development and implementation process. However, before a fully powered DCE can be conducted, a valid and reliable design must be created and estimates of β-coefficients are needed to inform sample size calculation.

Therefore, the aim of this study was to develop, and pilot test a DCE that evaluates key stakeholders’ preferences towards community exercise programmes for stroke survivors. The specific objectives are to: (1) develop a DCE design based on the available literature, and qualitative study of the target populations; (2) evaluate the feasibility of recruiting stroke stakeholders (stroke survivors, carers, clinicians, and fitness instructors) via online methods; (3) estimate β-coefficients to inform the sample size for the definitive DCE study.

## Methods

A DCE was developed to evaluate the preferences of four key stakeholders in post-stroke exercise services – stroke survivors, carers, clinicians, and fitness instructors. The feasibility of conducting a DCE with these populations was then tested. The study was conducted in accordance with the good research practices for conjoint analysis application in health from the International Society For Pharmacoeconomics and Outcomes Research (ISPOR) (Bridges *et al*., 2011). Ethical approval was granted from the Lancaster University Faculty of Health and Medicine Research Ethics Committee (FHM-2025-5726-RECR-2). All participants provided informed consent to access the DCE survey.

### DCE development

This study is part of a larger project aiming to co-produce a community-based exercise intervention for stroke survivors and their carers. The research question for this DCE was developed during a stakeholder panel meeting, during which members with expertise in health economics suggested the use of a DCE to elicit preferences of key stakeholders and evaluate if these differ between groups. After discussion, the panel agreed that the information generated from a DCE would be valuable for the intervention development process, and development of the DCE began.

### Attributes and levels

The selection of attributes and levels was informed by a variety of information sources: a scoping review of 66 included primary studies that investigated the experiences and perceptions of community-based exercise programmes for stroke survivors from the perspective of different stakeholders (Smith *et al*., 2026b); a series of focus groups with stroke survivors, carers, clinicians, and fitness instructors (Smith *et al*., 2026c); similar DCEs in the stroke literature (Jolliffe *et al*., 2024); and discussions with the intervention development stakeholder panel. Given the prevalence of cognitive impairment after stroke (Sun *et al*., 2014), efforts were made to reduce the cognitive burden of completing the DCE, and a maximum of six attributes (Table 1), eight choice tasks (per respondent), and two alternatives (plus opt-out) were used.

**Table 1.** Attributes and Levels.

| Attribute | Levels | Justification for inclusion |
| --- | --- | --- |
| Location | Home (online) | There is a growing number of telehealth interventions for stroke survivors, yet findings from the qualitative research suggest stroke survivors prefer to exercise outside of the home. There is also evidence that some stroke survivors do not want to engage with technology. Therefore, it is unclear if telehealth-based interventions are acceptable to stroke survivors. |
|  | Hybrid |  |
|  | Leisure centre |  |
| Price | £40/ month | Price is a commonly investigated attribute in the stroke literature and allows calculation of willingness to pay. The levels used represent the highest and lowest monthly costs of exercise referral schemes in England. |
|  | £20/ month |  |
|  | Free |  |
| Frequency | 1/ week | Findings from another stroke exercise DCE conflict with qualitative findings. It is unclear if stroke survivors prefer more, or less, frequent exercise sessions. The levels represent session frequencies available in exercise referral schemes in England. |
|  | 3/ week |  |
|  | 5/ week |  |
| Individualisation | Generic group exercises | The qualitative literature suggests individualisation is an important attribute to stroke survivors. However, providing greater levels of individualisation will result in greater costs to service providers, and therefore reduce feasibility. Calculating the willingness to pay for individualisation will be highly informative for service providers. |
|  | Targeted group exercises |  |
| Carer involvement | Carers not involved with the program | Carers are often overlooked during stroke rehabilitation, despite the importance of their role. The way in which carers should be engaged with exercise services is currently unknown. The levels used were identified from stakeholder consultations. |
|  | Carers join in with exercise sessions |  |
|  | Specific education and social sessions for carers |  |
| Recovery | 20% chance of meaningful improvement | Respondents were asked to consider what a meaningful improvement meant to them to ensure the outcome was individualised. Engagement with |
|  | 50% chance of meaningful improvement | higher intensity more efficient rehabilitation does not guarantee better outcomes but rather increases the chance of achieving them. A probability-based approach was used to better represent the unpredictability of the rehabilitation process. |
|  | 80% chance of meaningful improvement |  |

The six attributes utilised in the DCE design were: location, price, session frequency, individualisation, carer involvement, and recovery. Findings from Smith *et al*. (2026c) identified that stroke survivors may prefer to exercise at a leisure facility rather than at home, which contradicts findings from Geidl *et al*. (2018). There are a growing number of telehealth-based interventions for stroke survivors (Wu *et al*., 2022), yet it is unclear if stroke survivors want these services. Therefore, location was selected to investigate whether stroke stakeholders prefer to exercise at home, at a community venue, or a combination of both. Similarly for frequency, findings from Geidl *et al*. (2018) conflict with Smith *et al*. (2026c). Geidl *et al*. (2018) found a monotonic pattern for less frequent exercise sessions, whereas Smith *et al*. (2026c) found that stroke survivors would like more access to more frequent exercise sessions. Therefore, the direction of preference toward session frequency is currently unknown.

Price was included as it has been identified as a barrier to accessing community programmes (Smith *et al*., 2026b, 2026c), and allows calculation of willingness to pay. The levels used reflect current monthly prices of exercise referral schemes in England (Smith *et al*., 2026a).

Individualisation was a recurrent theme during literature reviews and stakeholder consultations (Smith *et al*., 2026b, 2026c), and has been identified as a dominant preference in the wider literature (Jolliffe *et al*., 2024). The calculation of willingness to pay for individualisation will be valuable for commissioners and intervention developers.

Carers are often overlooked in the rehabilitation process (Krishnan *et al*., 2017), despite clinical guidelines recommending that carers should receive information about stroke, be offered support, and be involved with rehabilitation activities (Sentinel Stroke National Audit Programme, 2023). Stakeholder consultations revealed that carers had varied attitudes towards their role in rehabilitation programmes, with some feeling they should not be involved, and others feeling carer-specific support should be provided (Smith *et al*., 2026c). Based on stakeholder consultations, the levels used were: carers not involved with the programme, carers join in with exercise sessions, and specific education and social sessions for carers.

Increased recovery has consistently been identified as a dominant preference by DCEs in the stroke rehabilitation literature (Jolliffe *et al*., 2024). This DCE used a slightly different paradigm using chance of meaningful improvement to better reflect the unpredictability of stroke rehabilitation. The use of a meaningful improvement makes the choice tasks more specific to the respondent’s needs and experiences, introducing a degree of individualisation.

### Choice tasks

Choice tasks were constructed using a Bayesian D-optimal design (Cook and Nachtrheim, 1980; Carlsson and Martinsson, 2003), in Stata MP version 18 (StataCorp LLC, 2023). A total of 16 choice sets were generated, which were divided into two blocks. Where appropriate, priors based on previous DCEs and the wider literature were used to inform the D-optimal design. Data for location and frequency was conflicting between sources, so a zero prior was used. The involvement of carers had not previously been investigated in a stroke DCE, and qualitative findings were conflicting, so a zero prior was used. Conservative priors were used for price (level 2: -0.1 and level 3: -0.5), individualisation (level 2: 0.15), and recovery (level 2: 0.2 and level 3: 0.7). Each choice task contained two alternatives, plus a neither option.

The choice tasks were incorporated into Qualtrics XM (Qualtrics, 2024), to allow online distribution. The online survey contained three sections: gaining informed consent, demographics, and DCE (Appendix H). Each of the four stakeholder types had specific demographics sections. After completing the demographics section, participants were provided with instructions for completing the DCE, then randomised to block A or block B. The survey was pre-tested by members of the intervention development stakeholder panel and deemed appropriate for the target populations. An example choice set, as presented in Qualtrics, is shown in Figure 1.

**Figure 1.**
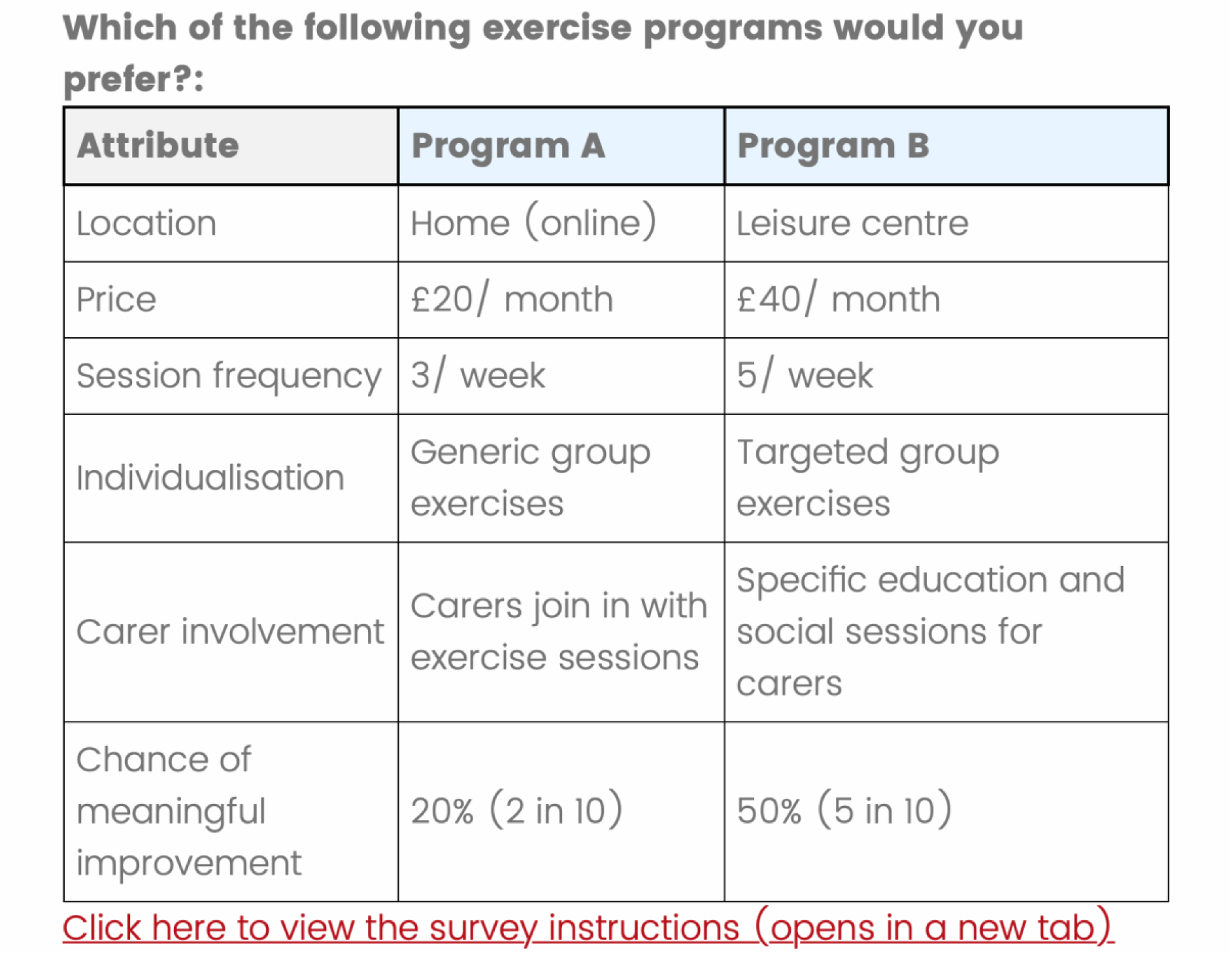
Example of Choice Task

### Data collection

Data were collected from stroke survivors, carers of stroke survivors, clinicians working with stroke survivors, and fitness instructors. There was no requirement for fitness instructors to have experience working with stroke survivors to be eligible. All participants were required to live in the UK and be aged 18 or over for inclusion. No restrictions were placed on stroke chronicity.

Stroke survivors and carers were recruited via convenience sampling of stroke support groups throughout the UK. Online searches of stroke support groups in each local authority in the UK were conducted to identify contact details for groups. The group admin was then contacted to invite their members to take part in the survey. A total of 377 local stroke support groups were contacted. Clinicians were recruited through social networks and two NHS trusts in North West England. Fitness instructors were recruited from a repository of contact details collected from a previous study (Smith *et al*., 2026a). No sample size target was set; however, to assess the feasibility of the online recruitment strategy, the aim was to recruit as many participants as possible using the above methods over a nine-month period.

### Data analysis

To assess the efficacy of the data collection methods, the characteristics of the respondents, drop-out rates, and completion times were described narratively.

Two separate models were developed for analysis – a main-effects model including all stakeholder types, and a stakeholder interaction model to investigate preference heterogeneity between stakeholder groups. For all models, price was coded as a continuous variable, to allow calculation of willingness to pay (McFadden, 1974), while all other variables were discrete. An alpha level of 0.05 was considered significant.

For the main effects analysis, both a conditional logit and mixed logit were initially developed. In the mixed logit model, preference heterogeneity was assessed using the estimated standard deviations of the random coefficients. A joint Wald test was used to test whether the standard deviations were jointly equal to zero. Due to evidence of significant preference heterogeneity, the conditional logit model was considered restrictive, as it does not account for preference heterogeneity (Hauber *et al*., 2016). Therefore, development was focussed on mixed logit model. When all attribute coefficients were specified as random, the mixed logit would converge when using 100 Halton draws but failed to converge when the number was increased to 500 and 1000. This suggested instability in the model, potentially reflecting empirical identification problems that were masked using a smaller number of draws (Chiou and Walker, 2007). Subsequently, the model was simplified by only allowing parameters with significant heterogeneity in the 100-draw mixed logit model to be random. The simplified model converged at 100, 500, and 1000 Halton draws, with very similar parameter estimates at 500 and 1000 draws.

Differences in preferences between stakeholder groups were investigated using a conditional logit model. Given the complexity (41 parameters) of the interaction model, it was not possible to use a mixed logit with the small pilot sample. As the analysis of individual stakeholder groups was expected to reduce heterogeneity, it was deemed appropriate to use a conditional logit for an exploratory analysis. Interactions were created between each attribute parameter and stakeholder group, with stroke survivors used as the reference group.

### Reliability and validity

To assess systematic alternative-specific preferences, an alternative-specific constant (ASC) was included in main-effects models. The ASC assesses respondents’ propensity to select a specific alternative (i.e., programme A or programme B), regardless of the underlying attributes and levels. To assess the content validity of the DCE design, the magnitude and direction of β-coefficients were compared to the priors used to inform the D-optimal design. Alignment of the priors and observed β-coefficients would suggest that respondents understand the context and content of the choice tasks.

### Sample size calculation

Sample size requirements for a future definitive DCE were estimated using the parametric approach proposed by de Bekker-Grob *et al*. (2015). The sample size calculation is based on a fixed-coefficient multinomial logit framework. Although the underlying theoretical approach can be extended to mixed logit models, this requires derivation of the asymptotic variance–covariance matrix under the specified mixed-logit model and experimental design. While a variance–covariance matrix can be obtained from the fitted pilot mixed logit model, this is conditional on the observed pilot data and is not directly equivalent to the prospective design-based matrix required for the calculation (Bliemer and Rose, 2010; de Bekker-Grob *et al*., 2015). As no directly applicable implementation was available, and no alternative calculations are available for mixed logit models, the published fixed-coefficient multinomial logit implementation was used. Subsequently, the resulting sample-size estimate relates to the detection of mean preference parameters rather than random-coefficient variance parameters. β-coefficients from the main effects model were used to inform the calculation. Minimum sample size thresholds were based on an alpha-level of 0.05 and 80% power. The output from the calculation was multiplied by two to reflect the blocked design of the DCE.

Given that the aim of this DCE is to assess preferences between stakeholder types, a sample size calculation should also be conducted for the stakeholder interaction analysis (de Bekker-Grob *et al*., 2015). However, there are currently no directly applicable parametric-based methods to calculate the minimum required sample size for the stakeholder interaction analysis. Because of this, two methods were used. First, the output from the main-effects sample size calculation was multiplied by four (for each of the stakeholder groups in the interaction model). Second, the Johnson and Orme (2003) rule of thumb approach was applied. The Johnson and Orme (2003) calculation uses the number of choice tasks completed by each respondent (*t*), the number of alternatives per choice task (*a*), and the largest number of analysis cells for any effect being estimated (*c*). For interaction effects, *c* was defined as the largest product of the number of levels involved in the interaction. The outputs from both methods are presented.

## Results

### Recruitment

Over the nine-month recruitment period, 377 unique stroke support groups across the UK were contacted. As the number of stroke support groups who shared the survey with their members is unknown, the true response rate cannot be calculated. However, the number of stroke survivor responses per support group contacted was 0.14, and for carers was 0.03.

Sixty-two emails were sent to fitness instructor teams, which generated 0.58 responses per email. Recruitment of clinicians was limited to two NHS trusts who shared the survey with staff, and a social media post, which produced 23 responses. However, only seven responses were from clinicians outside of North West England, suggesting most responses were from the NHS trusts.

### Survey completion

A total of 161 respondents started the survey, with 106 (66%) fully completing the survey. Twenty-nine (18%) respondents dropped-out before starting the DCE section, and 26 (16%) dropping-out during the DCE section. Of the 55 respondents who did not fully complete the survey, 15 (27%) were stroke survivors, 15 (27%) were fitness instructors, eight (15%) were clinicians, six (11%) were carers, and 11 (20%) did not specify their stakeholder type.

When excluding surveys that were completed in more than one session (as the timer continues between sessions), the median completion time was 8.57 minutes (Inter-quartile range: 5.50; 11.21). When stratified by stakeholder type, stroke survivors took the longest to complete the survey (median: 9.94 minutes), and clinicians took the least amount of time (median: 6.81).

### Participant characteristics

The characteristics of respondents, stratified by stakeholder type, are presented in Table 2. 132 participants completed at least one choice task, and therefore comprised the analytical sample. Of the 132 participants, 62 were stroke survivors (47%), 11 were carers (8%), 23 were clinicians (17%), and 36 were fitness instructors (27%). Stroke survivors were predominantly aged over 50, with most suffering their most recent stroke over six months ago (84.6%), almost half having a modified Rankin Scale score of 3, and 64% self-reporting cognitive impairment. All carers were female, and most were spouses/ partners of the stroke survivor. Clinicians were predominantly female and included a range of multidisciplinary professionals with varying levels of stroke-specific experience. Fitness instructors had diverse levels of experience, with most holding a Level 4 qualification. Across all participants, a larger proportion of females (n=88; 67%) completed the survey and most had access to a car (n=108; 82%). Respondents from North West England were also over-represented, with 42% of the sample living in the area. No responses from Wales or Northern Ireland were collected.

**Table 2.** Respondent characteristics.

|  |  |  | n | % |
| --- | --- | --- | --- | --- |
| Stroke survivors | Age | 31 - 40 | 3 | 4.84% |
|  |  | 41 - 50 | 8 | 12.90% |
|  |  | 51 - 60 | 13 | 20.97% |
|  |  | 61 - 70 | 21 | 33.87% |
|  |  | 71 - 80 | 14 | 22.58% |
|  |  | 81 - 90 | 3 | 4.84% |
|  | Gender | Male | 20 | 37.74% |
|  |  | Female | 33 | 62.26% |
|  | Time since stroke | 7 days - 3 months | 2 | 3.85% |
|  |  | 3 - 6 months | 6 | 11.54% |
|  |  | >6 months | 44 | 84.62% |
|  | mRS | 0 | 3 | 5.77% |
|  |  | 1 | 5 | 9.62% |
|  |  | 2 | 11 | 21.15% |
|  |  | 3 | 25 | 48.08% |
|  |  | 4 | 7 | 13.46% |
|  |  | 5 | 1 | 1.92% |
|  | Cognitive impairment | Yes | 32 | 64.00% |
|  |  | No | 18 | 36.00% |
|  | Employed | Yes | 13 | 25.00% |
|  |  | No | 39 | 75.00% |
|  | Financial strain | Extremely easy | 7 | 13.46% |
|  |  | Somewhat easy | 21 | 40.38% |
|  |  | Somewhat difficult | 14 | 26.92% |
|  |  | Extremely difficult | 5 | 9.62% |
|  |  | Prefer not to say | 5 | 9.62% |
|  | Access to car | Yes | 42 | 80.77% |
|  |  | No | 10 | 19.23% |
|  | Post-stroke exercise | Yes | 36 | 69.23% |
|  |  | No | 16 | 30.77% |
|  | Region | East Midlands | 1 | 1.96% |
|  |  | East of England | 3 | 5.88% |
|  |  | London | 2 | 3.92% |
|  |  | North East England | 2 | 3.92% |
|  |  | North West England | 32 | 62.75% |
|  |  | South East England | 4 | 7.84% |
|  |  | South West England | 1 | 1.96% |
|  |  | West Midlands | 1 | 1.96% |
|  |  | Yorkshire and the Humber | 2 | 3.92% |
|  |  | Scotland | 3 | 5.88% |
| Carers | Age | 31 - 40 | 1 | 9.09% |
|  |  | 41 - 50 | 0 | 0.00% |
|  |  | 51-60 | 2 | 18.18% |
|  |  | 61 - 70 | 1 | 9.09% |
|  |  | 71 - 80 | 5 | 45.45% |
|  |  | 81 - 90 | 2 | 18.18% |
|  | Gender | Male | 0 | 0.00% |
|  |  | Female | 11 | 100.00% |
|  | Time in role | 6 - 12 months | 2 | 18.18% |
|  |  | 1 - 5 years | 3 | 27.27% |
|  |  | 6 - 10 years | 3 | 27.27% |
|  |  | >10 years | 3 | 27.27% |
|  | Relation to stroke survivor | Spouse/ partner | 9 | 81.82% |
|  |  | Friend | 1 | 9.09% |
|  |  | Volunteer | 1 | 9.09% |
|  | Employed | Yes | 2 | 18.18% |
|  |  | No | 9 | 81.82% |
|  | Carer strain | None at all | 1 | 9.09% |
|  |  | A little | 2 | 18.18% |
|  |  | A moderate amount | 2 | 18.18% |
|  |  | A great deal | 4 | 36.36% |
|  |  | A lot | 2 | 18.18% |
|  | Car access | Yes | 10 | 90.91% |
|  |  | No | 1 | 9.09% |
|  | Financial strain | Extremely easy | 1 | 9.09% |
|  |  | Somewhat easy | 3 | 27.27% |
|  |  | Somewhat difficult | 3 | 27.27% |
|  |  | Extremely difficult | 0 | 0.00% |
|  |  | Prefer not to say | 4 | 36.36% |
|  | Region | East Midlands | 0 | 0.00% |
|  |  | East of England | 2 | 18.18% |
|  |  | London | 0 | 0.00% |
|  |  | North East England | 0 | 0.00% |
|  |  | North West England | 7 | 63.64% |
|  |  | South East England | 0 | 0.00% |
|  |  | South West England | 0 | 0.00% |
|  |  | West Midlands | 2 | 18.18% |
|  |  | Yorkshire and the Humber | 0 | 0.00% |
|  |  | Scotland | 0 | 0.00% |
| Clinicians | Age | 18 - 30 | 5 | 21.74% |
|  |  | 31 - 40 | 8 | 34.78% |
|  |  | 41 - 50 | 5 | 21.74% |
|  |  | 51 - 60 | 4 | 17.39% |
|  |  | 61 - 70 | 1 | 4.35% |
|  | Gender | Male | 4 | 17.39% |
|  |  | Female | 19 | 82.61% |
|  | Profession | Physiotherapist | 7 | 30.43% |
|  |  | Occupational therapist | 6 | 26.09% |
|  |  | SALT | 3 | 13.04% |
|  |  | Therapy assistant | 2 | 8.70% |
|  |  | Physician | 1 | 4.35% |
|  |  | Clinical psychologist | 1 | 4.35% |
|  |  | Other | 3 | 13.04% |
|  | Stroke-specific experience | 1 - 5 years | 8 | 34.78% |
|  |  | 6 - 10 years | 6 | 26.09% |
|  |  | 11 - 15 years | 6 | 26.09% |
|  |  | 16 - 20 years | 1 | 4.35% |
|  |  | >20 years | 2 | 8.70% |
|  | Car access | Yes | 21 | 91.30% |
|  |  | No | 1 | 4.35% |
|  |  | Prefer not to say | 1 | 4.35% |
|  | Region | East Midlands | 1 | 4.35% |
|  |  | East of England | 0 | 0.00% |
|  |  | London | 0 | 0.00% |
|  |  | North East England | 3 | 13.04% |
|  |  | North West England | 16 | 69.57% |
|  |  | South East England | 1 | 4.35% |
|  |  | South West England | 0 | 0.00% |
|  |  | West Midlands | 2 | 8.70% |
|  |  | Yorkshire and the Humber | 0 | 0.00% |
|  |  | Scotland | 0 | 0.00% |
| Fitness instructors | Age | 18 - 30 | 7 | 19.44% |
|  |  | 31 - 40 | 9 | 25.00% |
|  |  | 41 - 50 | 5 | 13.89% |
|  |  | 51 - 60 | 12 | 33.33% |
|  |  | 61 - 70 | 3 | 8.33% |
|  | Gender | Male | 11 | 30.56% |
|  |  | Female | 25 | 69.44% |
|  | Experience | 1 - 5 years | 10 | 27.78% |
|  |  | 6 - 10 years | 6 | 16.67% |
|  |  | 11 - 15 years | 9 | 25.00% |
|  |  | 16 - 20 years | 3 | 8.33% |
|  |  | >20 years | 8 | 22.22% |
|  | Highest qualification | Level 2 | 5 | 13.89% |
|  |  | Level 3 | 6 | 16.67% |
|  |  | Level 4 | 22 | 61.11% |
|  |  | Above level 4 | 2 | 5.56% |
|  |  | Prefer not to say | 1 | 2.78% |
|  | Exposure to stroke survivors | None at all | 3 | 8.33% |
|  |  | Limited | 16 | 44.44% |
|  |  | Moderate | 13 | 36.11% |
|  |  | Extensive | 4 | 11.11% |
|  | Car access | Yes | 35 | 97.22% |
|  |  | No | 1 | 2.78% |
|  | Region | East Midlands | 7 | 19.44% |
|  |  | East of England | 6 | 16.67% |
|  |  | London | 1 | 2.78% |
|  |  | North East England | 7 | 19.44% |
|  |  | North West England | 0 | 0.00% |
|  |  | South East England | 8 | 22.22% |
|  |  | South West England | 4 | 11.11% |
|  |  | West Midlands | 2 | 5.56% |
|  |  | Yorkshire and the Humber | 1 | 2.78% |
|  |  | Scotland | 0 | 0.00% |

### Stakeholder preferences

#### Main effects

During development of the mixed logit model, five parameters demonstrated evidence of significant preference heterogeneity – location (hybrid and leisure centre), frequency (five sessions/ week), carer involvement (carer-specific sessions), recovery (50% chance), and recovery (80% chance). Subsequently, these parameters were specified as random, and all others specified as fixed, in the final mixed logit model. The findings from the main-effects mixed logit model are presented in Table 3. The results show that stroke stakeholders have a significant preference for exercise programmes that: take place in a leisure centre, have lower price, are individualised, have carer-specific sessions, and offer greater chance of meaningful recovery. No significant preference was identified for session frequency, although estimates increased with session frequency. Attributes followed the expected monotonic pattern, that aligned with the priors used in the DCE design. A joint Wald test of the random-coefficient standard deviations indicated evidence of preference heterogeneity (χ²(5) = 35.37, p<0.001). The ASC did not produce a statistically significant alternative-specific preference.

**Table 3.**
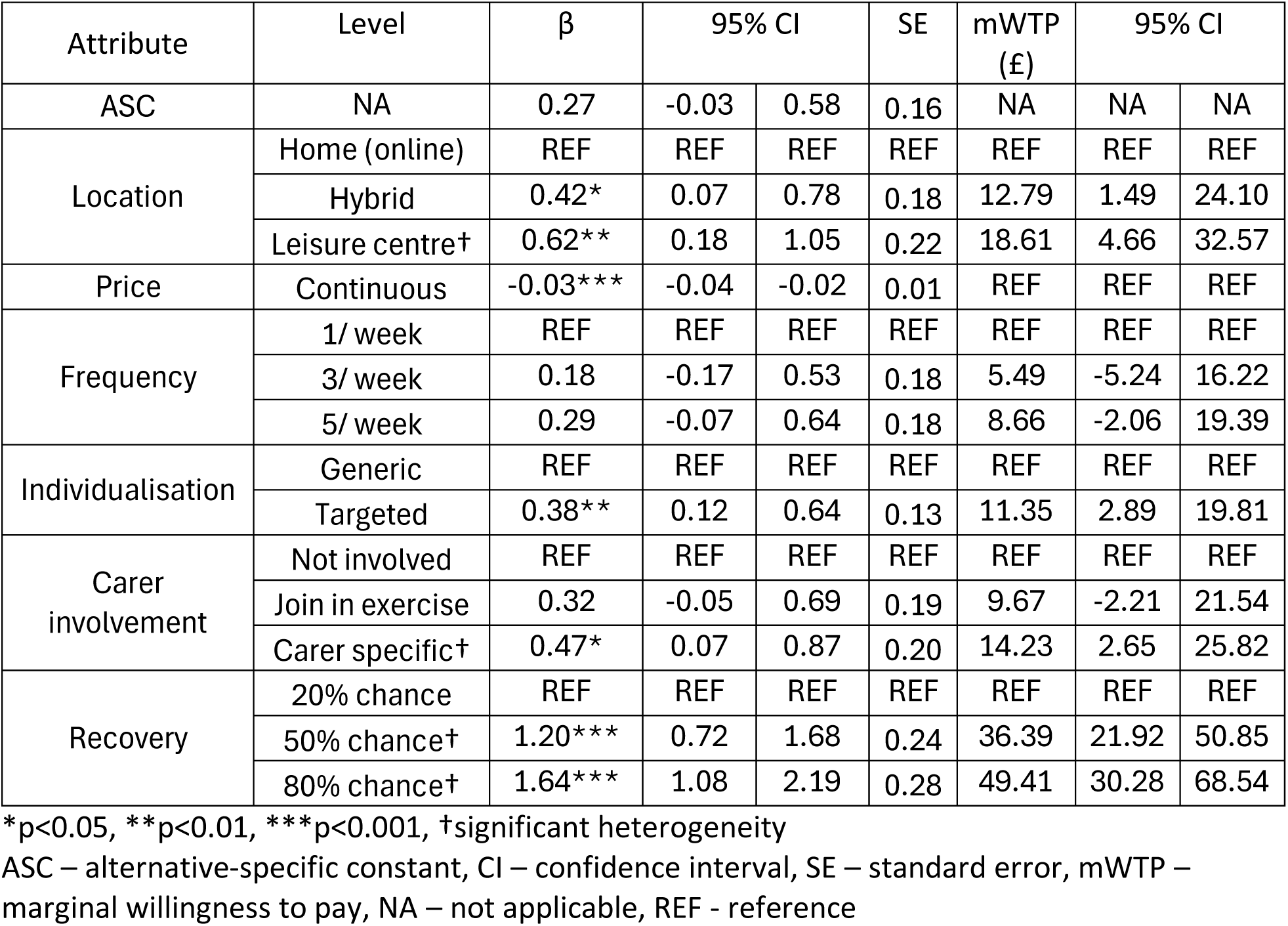
Results from main-effects mixed logit model.

| Attribute | Level | $\beta$ | 95% CI | | SE | mWTP (£) | 95% CI | |
| --- | --- | --- | --- | --- | --- | --- | --- | --- |
| ASC | NA | 0.27 | -0.03 | 0.58 | 0.16 | NA | NA | NA |
| Location | Home (online) | REF | REF | REF | REF | REF | REF | REF |
|  | Hybrid | 0.42* | 0.07 | 0.78 | 0.18 | 12.79 | 1.49 | 24.10 |
|  | Leisure centre† | 0.62** | 0.18 | 1.05 | 0.22 | 18.61 | 4.66 | 32.57 |
| Price | Continuous | -0.03*** | -0.04 | -0.02 | 0.01 | REF | REF | REF |
| Frequency | 1/ week | REF | REF | REF | REF | REF | REF | REF |
|  | 3/ week | 0.18 | -0.17 | 0.53 | 0.18 | 5.49 | -5.24 | 16.22 |
|  | 5/ week | 0.29 | -0.07 | 0.64 | 0.18 | 8.66 | -2.06 | 19.39 |
| Individualisation | Generic | REF | REF | REF | REF | REF | REF | REF |
|  | Targeted | 0.38** | 0.12 | 0.64 | 0.13 | 11.35 | 2.89 | 19.81 |
| Carer involvement | Not involved | REF | REF | REF | REF | REF | REF | REF |
|  | Join in exercise | 0.32 | -0.05 | 0.69 | 0.19 | 9.67 | -2.21 | 21.54 |
|  | Carer specific† | 0.47* | 0.07 | 0.87 | 0.20 | 14.23 | 2.65 | 25.82 |
| Recovery | 20% chance | REF | REF | REF | REF | REF | REF | REF |
|  | 50% chance† | 1.20*** | 0.72 | 1.68 | 0.24 | 36.39 | 21.92 | 50.85 |
|  | 80% chance† | 1.64*** | 1.08 | 2.19 | 0.28 | 49.41 | 30.28 | 68.54 |

\*p<0.05, \*\*p<0.01, \*\*\*p<0.001, †significant heterogeneity
ASC – alternative-specific constant, CI – confidence interval, SE – standard error, mWTP – marginal willingness to pay, NA – not applicable, REF - reference

Marginal willingness-to-pay (mWTP) estimates are presented in Figure 2. The largest mWTP estimates were observed for improvements in chance of recovery. Respondents were willing to pay an additional £49.41 per month for an 80% chance of meaningful recovery compared to a 20% chance. Respondents were also willing to pay an additional £18.61 per month to attend an exercise programme at a leisure centre compared to a home-based online programme. Individualisation was valued at an additional £11.35 per month compared to a generic programme, while carer-specific sessions were valued at an additional £14.23 per month, compared with no carer involvement. The mWTP estimates for session frequency were uncertain, with confidence intervals crossing zero, although estimates increased with greater session frequency.

**Figure 2.**
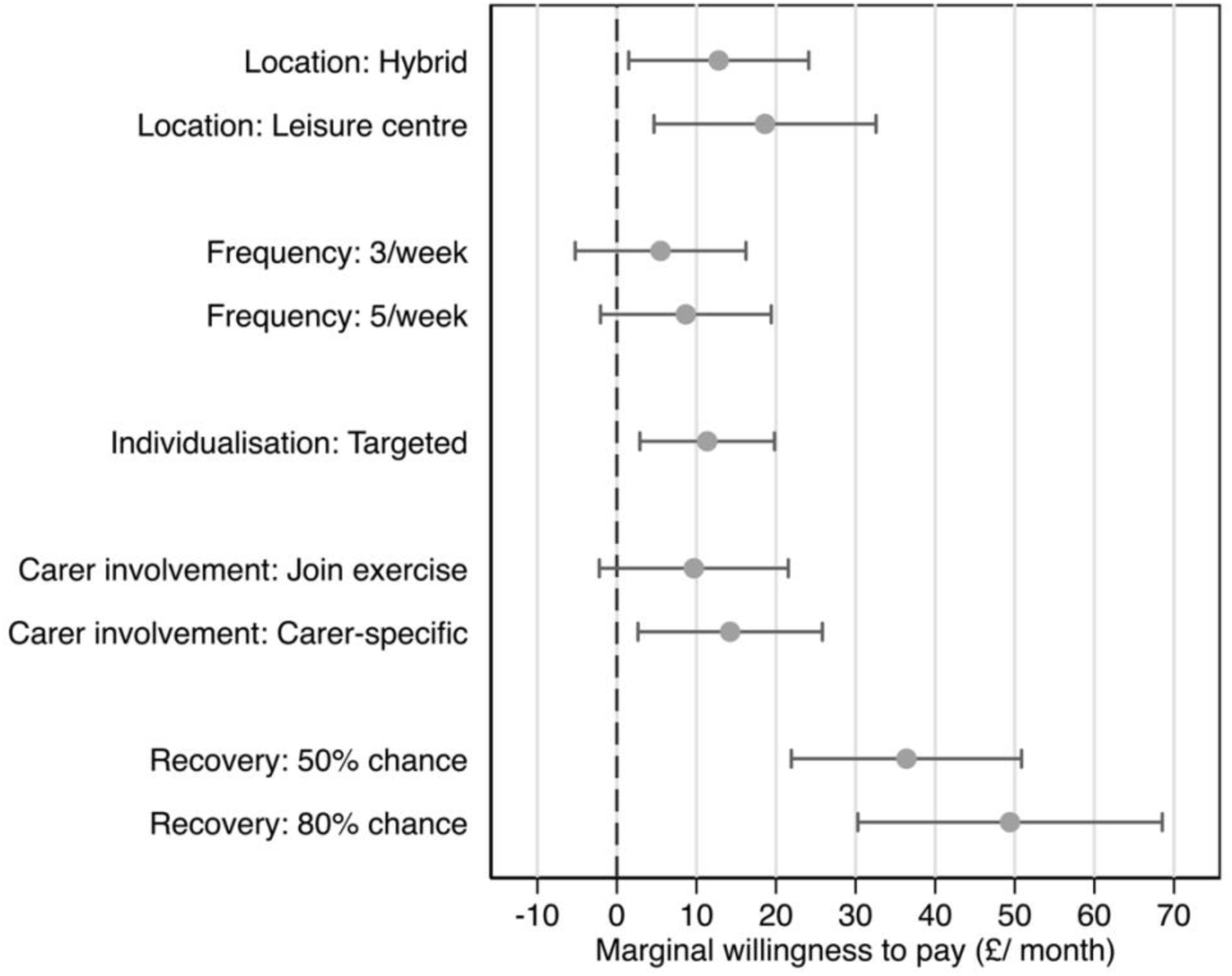
Marginal willingness-to-pay.

#### Stakeholder interactions

The findings from the stakeholder interaction conditional logit model are presented in Table 4. The results from this model should be considered exploratory. Significant differences in preferences were found between stroke survivors and clinicians for price, carer involvement and recovery. Compared to stroke survivors, clinicians were more price sensitive, demonstrated stronger preference for carer-specific sessions, and placed greater importance on higher chance of recovery.

**Table 4.**
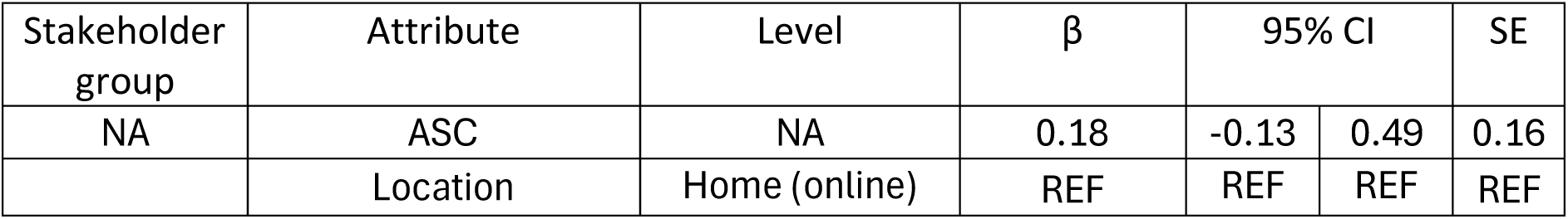
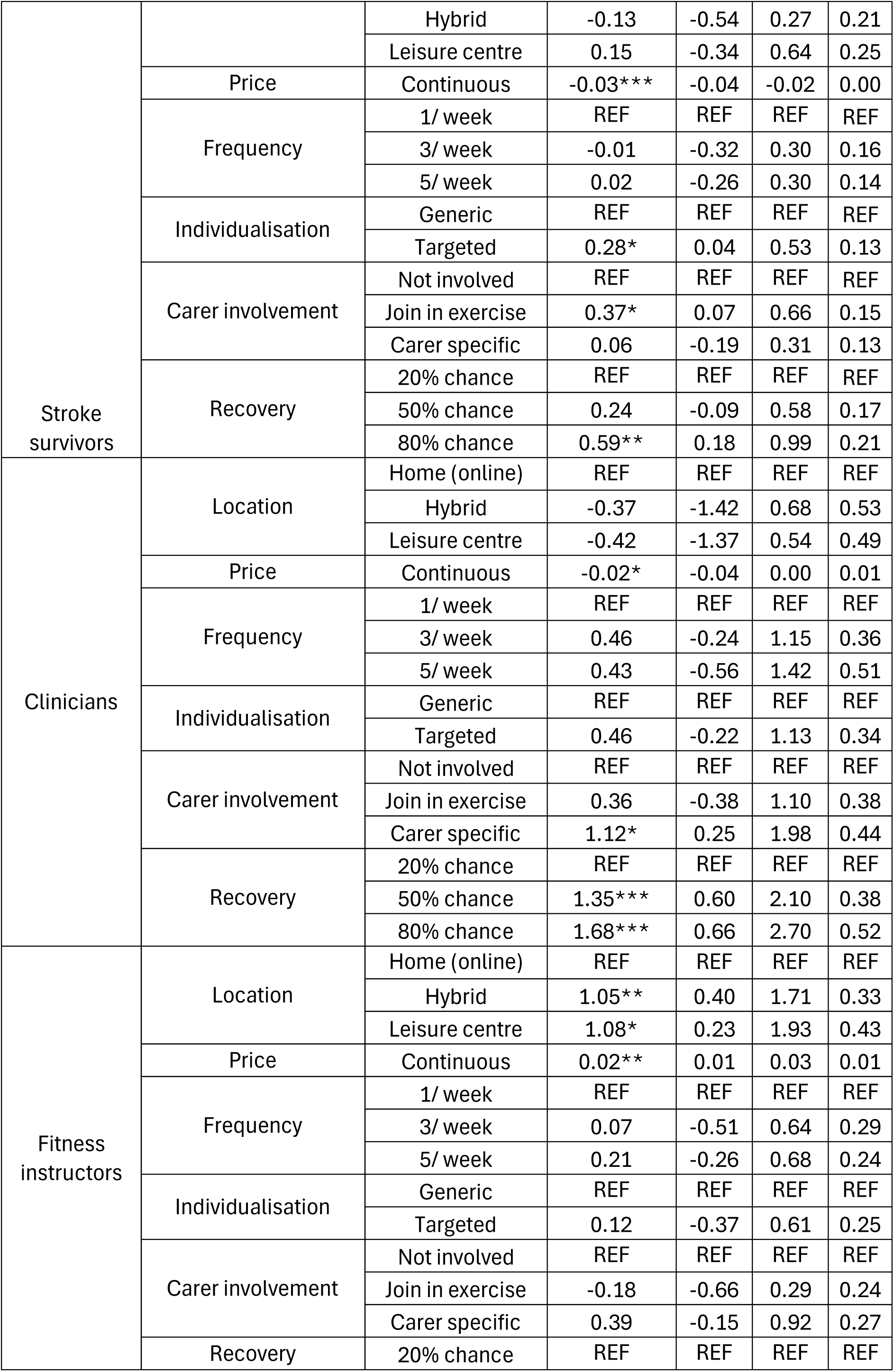
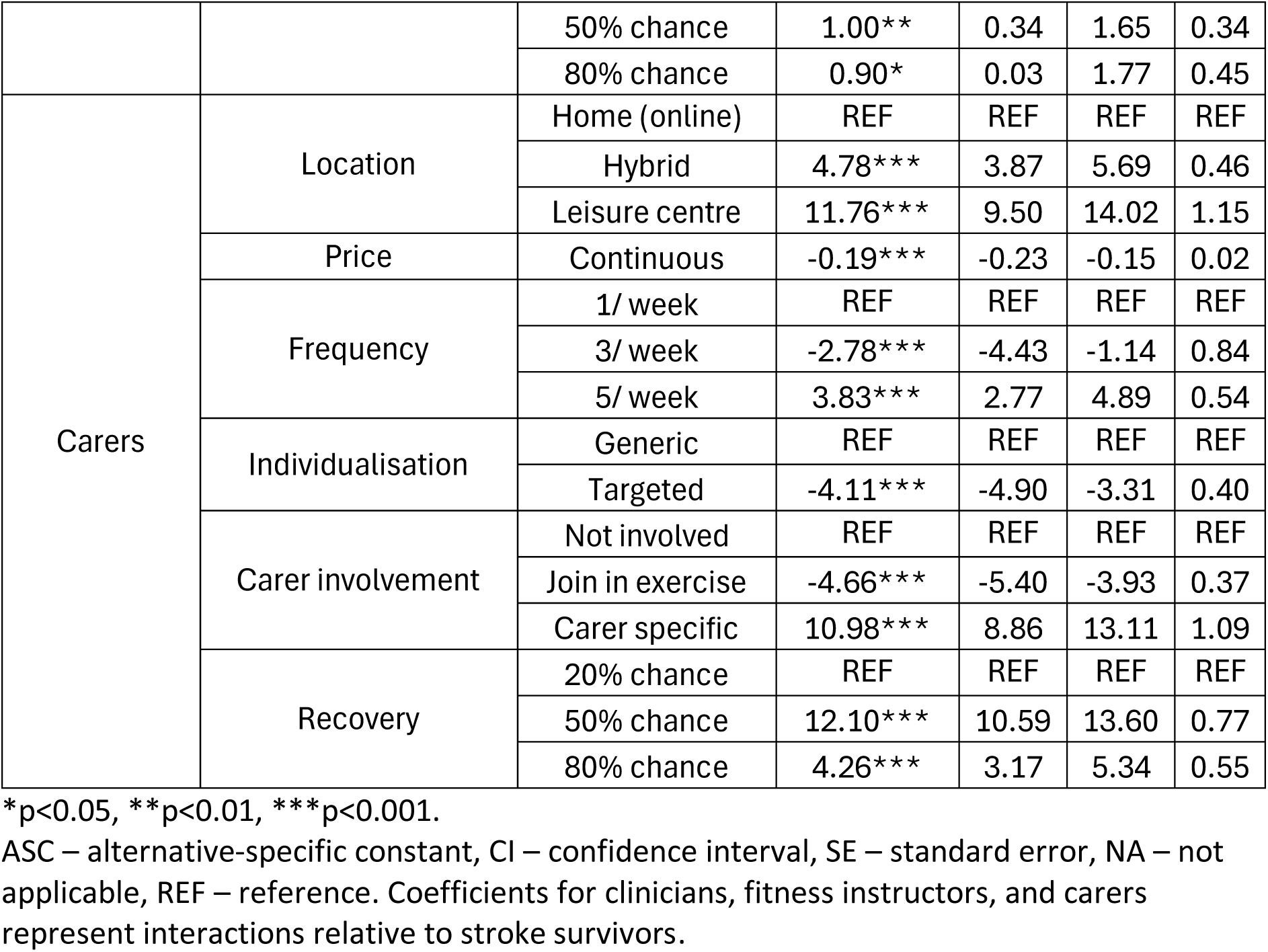
Results from subgroup interaction model.

Significant differences were also found between stroke survivors and fitness instructors. Compared to stroke survivors, fitness instructors demonstrated stronger preference for hybrid and leisure centre delivery and placed greater importance on higher chance of recovery. Stroke survivors were more price sensitive than fitness instructors.

Due to the low number of carer responses, carer interaction estimates were characterised by extremely large β-coefficients, with significant variation in both magnitude and direction across attribute levels. These estimates were considered unstable and were not used to draw conclusions regarding carer preferences.

#### Sample size estimation

Using the de Bekker-Grob *et al*. (2015) parametric approach, the estimated sample size requirements varied across parameters in the main-effects model (Table 5). The largest sample size requirement was observed for session frequency (three sessions per week), corresponding to a minimum sample of 334, which was driven by the relatively small β-coefficient. The next largest sample size requirement was for carer involvement (join in exercise), corresponding to a minimum sample of 118.

**Table 5.** Main effects sample size estimation.

| Attribute | Level | Estimated sample |
| --- | --- | --- |
| Location | Hybrid | 80 |
|  | Leisure centre | 23 |
| Price | Continuous | 8 |
| Frequency | 3 sessions / week | 334 |
|  | 5 sessions / week | 116 |
| Individualisation | Targeted | 52 |
| Carer involvement | Join in exercise | 118 |
|  | Specific sessions | 51 |
| Recovery | 50% chance | 8 |
|  | 80% chance | 5 |

For the stakeholder interaction model, the estimate of 118 participants from the main-effects calculation was multiplied by four, producing an estimate of 472 participants (assuming equal recruitment between groups). The estimate of 334 participants was not used, as this was driven by a small coefficient that may not represent a meaningful preference. The Johnson and Orme (2003) rule of thumb produced an indicative sample size of 252 participants, or 63 participants per stakeholder group.

## Discussion

This pilot DCE provides preliminary evidence of the preferences of stroke stakeholders towards exercise programmes. Generally, price and recovery had the strongest utilities, but the relative magnitude of these differ across stakeholder types. Stroke survivors placed less importance on the amount of recovery achieved compared to clinicians and fitness instructors. Clinicians were the most price sensitive stakeholder group, followed by stroke survivors, then fitness instructors. As this is a pilot study with a relatively low sample size, these results should be interpreted cautiously but justify further investigation in a larger scale study.

The DCE design piloted in this study was found to be robust and suitable for large-scale testing. The estimated β-coefficients for each attribute aligned to *a priori* expectations and each level demonstrated a monotonic pattern, suggesting the choice tasks were well understood by the respondents (Pearce *et al*., 2021), and levels were sufficiently spaced to produce meaningful differences to stakeholder decision making. The findings from this study suggest that the attributes and levels used are important components of exercise programmes for stroke survivors. The number of attributes used in the DCE design was deemed appropriate based on the time taken to complete the survey. Stroke survivors took the longest to complete the survey (median: 9.94 minutes), which is considerably shorter than other DCEs with stroke survivors (Firth *et al*., 2026), suggesting the survey design is not overly burdensome for respondents. Based on the findings of this pilot, all attributes and levels should be carried forward to a larger-scale study.

The findings from this pilot DCE can be compared with Geidl *et al*. (2018), who conducted a similar DCE with 103 stroke survivors undergoing inpatient rehabilitation. Geidl *et al*. (2018) found no significant preference for exercise location. In contrast, the main-effects analysis in the present study identified significant preferences for both hybrid, and leisure centre delivery compared to home-based delivery, which aligned with preliminary qualitative findings (Smith *et al*., 2026c). However, this difference may be explained by the inclusion of clinicians, fitness instructors, and carers in the mixed-effect model, as the stakeholder interaction analysis found stroke survivors had no significant preference for location. Geidl *et al*. (2018) also found no significant preference towards session frequency, which is consistent with the present findings. However, the direction of coefficients were conflicting, with Geidl *et al*. (2018) finding higher coefficients for lower session frequency. Previous qualitative findings support the direction found in this present study (Smith *et al*., 2026c). A lack of statistical power may have contributed to the absence of statistically significant frequency effects. Although Geidl *et al*. (2018) reports that the analysis was fully powered, a rule-of-thumb sample size calculation was used, which does not account for anticipated coefficients and may therefore provide less informative sample-size estimates than parametric approaches (de Bekker-Grob *et al*., 2015). Overall, these findings suggest that preferences for programme location may differ across stakeholder types, while preferences for session frequency remain uncertain and warrant further investigation in a larger, adequately powered DCE.

Geidl *et al*. (2018) acknowledged that the use of a fixed logit model was a limitation of the study, as it cannot account for preference heterogeneity. Although the present study investigated preference heterogeneity between stakeholder groups, heterogeneity within stakeholder groups has not yet been investigated in the context of post-stroke exercise programmes. The conflicting direction of coefficients found for session frequency suggest that preferences may vary between stroke populations, potentially due to factors such as stroke chronicity. While sub-group heterogeneity could not be assessed in this pilot due to the small sample size, information was collected in the demographics section that would facilitate the investigation of sub-group preference heterogeneity in a larger-scale study. The time taken to complete the survey was considerably shorter than other DCEs in the stroke literature, likely due to the lower number of choice tasks (Firth *et al*., 2026); however, the completion rate in the present study was lower than the average of 82.3% in health-related DCEs (Nouwens *et al*., 2025). Cognitive impairment is a common problem after stroke (Sun *et al*., 2014), and minimising cognitive burden associated with completing DCEs may be important for reducing drop-out. Future larger-scale studies should collect sufficient demographic and clinical information to investigate intragroup preference heterogeneity, while avoiding excessive data collection that may increase cognitive burden and therefore reduce completion rates.

DCEs in the stroke literature have consistently reported price and recovery as attributes with high utility (Firth *et al*., 2026; Doshi *et al*., 2023; Noto *et al*., 2023; Jolliffe *et al*., 2024), which is consistent with this pilot study. However, this study goes further by investigating differences between stakeholders. Although other DCEs in the stroke literature have included multiple stakeholders (Burton *et al*., 2014; Laver *et al*., 2013), none have used stakeholder group as an interaction to directly compare preferences. The stakeholder interaction model found that preferences toward recovery differed significantly between stakeholder groups, with both fitness instructors and clinicians having stronger preferences for programmes that offer greater chance of recovery. This may partly be explained by the disability paradox, where people living with a disability rate their quality of life more positively than people without lived experience expect (Albrecht and Devlieger, 1999).

Hence, respondents without stroke-related impairments may focus more on the recovery aspect, whereas stroke survivors give more consideration to factors related to convenience and burden. This is most evident when comparing preferences between stroke survivors and fitness instructors, who likely have less awareness of stroke-related impairments than clinicians, with stroke survivors being significantly more price sensitive, and having less preference for hybrid and leisure centre delivery, which may reflect the burden of transporting to a community venue.

Problems with the online recruitment strategy were identified, and changes will be needed to recruit for a fully powered study. Recruitment of carers was particularly challenging, with only 11 carer responses recorded from 377 stroke support group contacts. Recruitment of stroke survivors was also inefficient using this method. For the final study, in-person recruitment through the NHS should be used to recruit an adequate number of stroke survivors and carers. Another advantage of NHS recruitment is that it allows recruitment of more stroke survivors in the acute and sub-acute phases, who were under-represented in this pilot. The recruitment of clinicians was also a challenge, which is a prevalent problem in the survey literature (Klabunde *et al*., 2013). Links with other NHS trusts will be needed to effectively recruit clinicians, as social media recruitment proved to be ineffective. Utilisation of professional bodies, such as the Chartered Society of Physiotherapists and Royal College of Occupational Therapists, could also be used to facilitate recruitment of clinicians. The recruitment of fitness instructors was the most efficient, with 36 responses gained from 62 email contacts. However, these were contacts who were happy to be contacted for research opportunities, and response rates for other leisure centres may be lower. Nonetheless, as there are 1,419 leisure centres in the UK (Sport England, 2025), the online method of recruitment can be scaled up to achieve adequate sampling. The information gathered from this pilot study will allow recruitment strategies to be effectively modified for a fully powered study.

The findings of this pilot study provide guidance for sample size requirements for a future definitive DCE. The estimated requirement of 334 respondents for the main-effects model was driven by the relatively small coefficient for three sessions per week, and it is unclear if an effect of this magnitude would be sufficiently meaningful to justify powering a study to detect it (Lakens, 2022). The sample size requirement for the next smallest parameter was substantially lower, at 118 participants, which may represent a more meaningful estimate. However, as the main aim of the DCE is to identify differences in preference between stakeholder groups, a future study should be primarily powered for the stakeholder interaction analysis. As there are no directly applicable parametric-based approaches for the stakeholder interaction model, a pragmatic approach was used, in which the output of the main effects sample size calculation was multiplied by four, to represent each stakeholder group. This provided a sample size estimate of 472 participants, which is close to the median sample of 394 in health-related DCEs (Nouwens *et al*., 2025). Although rule-of-thumb approaches have been criticised for their lack of accuracy (de Bekker-Grob *et al*., 2015), the Johnson and Orme (2003) calculation was also used, due to the sparsity of alternative approaches, which produced a considerably lower estimate of 252 respondents. As the Johnson and Orme (2003) calculation does not account for effect sizes or statistical power, this lower estimate should be interpreted with extreme caution. The findings suggest that a larger sample size is required to investigate stakeholder interactions, and a target of 252 respondents may be a useful lower planning threshold, while a larger sample of 472 would provide greater precision.

### Strengths and limitations

This study has several strengths. First, the DCE considered perspectives from four key stakeholder groups who would receive, support, or deliver the proposed intervention, enabling preferences to be examined from both service user and provider perspectives. The planned main-effects and stakeholder interaction analyses were successfully tested on the pilot data, and findings were used to inform the sample size for a future definitive DCE. Furthermore, the feasibility of recruiting a diverse range of stakeholders using online methods was evaluated, providing important considerations for future studies. The DCE design was informed by thorough reviews of the literature, stakeholder consultations, and evaluations of currently available exercise services, which resulted in an effective DCE design. This pilot demonstrated the feasibility of the DCE design and analytical approach, supporting its use in a larger-scale study. However, some limitations should be considered. The recruitment strategy was inefficient and resulted in uneven recruitment between stakeholder groups, particularly among carers. Subsequently, stakeholder interaction estimates were imprecise, with substantial instability observed in the carer estimates. The recruitment strategy failed to recruit acute stroke survivors, which may have limited the range of preferences represented in the pilot sample. The lower completion rate compared with other health-related DCEs suggests that respondent burden should be considered when refining the survey for a definitive study. Finally, a formal power calculation could not be performed for the stakeholder interaction analysis, and the results only provide estimates of sample size requirements.

### Conclusions

This pilot has demonstrated the feasibility of a DCE aiming to investigate differences in preferences towards post-stroke exercise programmes in four key stakeholders. The DCE design used was found to be reliable, and the analytical approach appropriate. However, changes to the recruitment strategy will be needed to recruit for a fully-powered study, as the online recruitment strategy employed in this pilot was ineffective. More consideration needs to be given to recruiting carers and clinicians, to ensure a more even distribution of stakeholders for the final study.

Despite the exploratory nature of the study, the mWTP estimates provide valuable data for those developing exercise interventions for stroke survivors. Stroke stakeholders place the greatest value on the chance of making a meaningful recovery, leisure centre delivery, carer-specific sessions, and individualisation. A fully-powered DCE will provide more precise estimates to support the development and economic evaluations of future exercise-based interventions for stroke.

## Declarations

### Ethics approval and consent to participate

Ethical approval was granted by the Lancaster University Faculty of Health and Medicine Research Ethics Committee (FHM-2025-5726-RECR-2). All participants provided informed consent to participate in the study. The study was conducted in accordance with the Declaration of Helsinki.

### Availability of data and materials

All data generated by this study is available upon reasonable request to the corresponding author.

## Authors contributions

Conceptualisation - JS

Methodology – JS, LF

Software - JS

Formal analysis – JS, LF

Investigation - JS

Data curation - JS

Writing (original draft) - JS

Writing (review & editing) - PN, HJ, LF

Visualisation - JS

Supervision - PN, CH, HJ, LF

## Supporting information

Appendix H - Online survey

## Data Availability

All data produced in the present study are available upon reasonable request to the authors.

## Acknowledgements

Not applicable

