## Appendix H - Online survey for "Developing and piloting a discrete choice experiment to assess stakeholder preferences for community-based exercise programmes after stroke"

Final DCE survey design

Online survey preview: <https://lancasteruni.eu.qualtrics.com/jfe/preview/previewId/9fee485a-ab2d-4d0f-b50d-e7aafef3751a/SV_01xHXxmaF3VA9tc?Q_CHL=preview&Q_SurveyVersionID=current>

Two of the questions in this survey (Q38 and Q46) are written in HTML and will only be displayed with the correct formatting in the above survey link. These two questions use the loop and merge function, which repeats the same question design, with different contents in the table eight times. The eight repeats are the same for each block (i.e., everyone randomized to block A will answer the same 8 questions).

To move through the survey preview, you will have to complete some of the questions to move onto the next section. As this is a preview, your responses will not be recorded.

Survey Flow

Standard: Consent (1 Question)

Block: Identify stakeholder type (1 Question)

Branch: New Branch

If

If Are you a... Stroke survivor Is Selected

Standard: Stroke survivor demographics (10 Questions)

Branch: New Branch

If

If Are you a... Carer of a stroke survivor Is Selected

Standard: Carer demographics  (10 Questions)

Branch: New Branch

If

If Are you a... Clinician Is Selected

Standard: Clinician demographics  (6 Questions)

Branch: New Branch

If

If Are you a... Fitness instructor Is Selected

Block: Fitness instructor demographics (6 Questions)

Block: Survey instructions (2 Questions)

BlockRandomizer: 1 - Evenly Present Elements

Standard: DCE block 1 (2 Questions)

Standard: DCE block 2 (2 Questions)

Standard: Future research (2 Questions)

| Page Break |
| --- |

Start of Block: Consent

Q43 Please read the following statements: 1. I confirm that I have read the information sheet and fully understand what is expected of me within this study	Click here to view the information sheet (opens in a new tab)	2. I confirm that I have had the opportunity to ask any questions and to have them answered. 3. I understand that my participation is voluntary and that I am free to withdraw at any time without giving any reason. 4. I understand that some questions may ask about special category data and that I have the option of not answering them. 5. I understand that once my data have been anonymised it might not be possible for it to be withdrawn. 6. I understand that the information from my survey will be pooled with other participants’ responses, anonymised and may be published. 7. I consent to take part in the above survey. Having read the participant information sheet and the above statements, do you consent to take part in the survey?

- Yes (1)
- No (2)

Skip To: End of Survey If Please read the following statements: 1. I confirm that I have read the information sheet and ful... = No

End of Block: Consent

Start of Block: Identify stakeholder type

Q1 Are you a...

- Stroke survivor (1)
- Carer of a stroke survivor (2)
- Clinician (3)
- Fitness instructor (4)

End of Block: Identify stakeholder type

Start of Block: Stroke survivor demographics

Q2 What is your age?

- 18 - 30 (4)
- 31 - 40 (5)
- 41 - 50 (6)
- 51 - 60 (7)
- 61 - 70 (8)
- 71 - 80 (9)
- 81 - 90 (10)
- 91 or older (11)
- Prefer not to say (12)

Q11 What is your gender?

- Male (1)
- Female (2)
- Prefer to self-describe (3) __________________________________________________
- Prefer not to say (4)

Q4 How long ago was your most recent stroke?

- Less than 7 days ago (1)
- More than 7 days but less than 3 months ago (2)
- More than 3 months ago but less than 6 months ago (3)
- Longer than 6 months ago (4)
- Prefer not to say (5)

Q5 Which of the following options best describes your current level of disability?

- No disability at all (1)
- No significant disability despite symptoms; able to carry out all usual duties and activities (2)
- Slight disability; unable to carry out all previous activities, but able to look after own affairs without assistance (3)
- Moderate disability; requiring some help, but able to walk without assistance (4)
- Moderately severe disability; unable to walk and attend to bodily needs without assistance (5)
- Severe disability; bedridden, incontinent and requiring constant nursing care and attention (6)
- Prefer not to say (7)

Q8 Do you currently experience difficulty with your thinking skills (memory, problem solving, attention)?

- No (3)
- Yes (4)
- Prefer not to say (5)

Q6 Do you or someone you live with have access to a car?

- Yes (1)
- No (2)
- Prefer not to say (3)

Q35 Are you currently employed?

- No (3)
- Yes (4)
- Prefer not to say (5)

Q7 How easy or difficult is it for your household to make ends meet financially each month?

- Extremely difficult (1)
- Somewhat difficult (2)
- Somewhat easy (3)
- Extremely easy (4)
- Prefer not to say (5)

Q9 Have you taken part in any exercise programs since having a stroke?

- No (3)
- Yes (4)
- Prefer not to say (5)

Q25 What region do you currently live in?

- North East England (1)
- North West England (2)
- Yorkshire and the Humber (3)
- East Midlands (4)
- West Midlands (5)
- East of England (6)
- London (7)
- South East England (8)
- South West England (9)
- Scotland (10)
- Wales (11)
- Northern Ireland (12)
- Prefer not to say (13)

End of Block: Stroke survivor demographics

Start of Block: Carer demographics

Q43 What is your age?

- 18 - 30 (4)
- 31 - 40 (5)
- 41 - 50 (6)
- 51 - 60 (7)
- 61 - 70 (8)
- 71 - 80 (9)
- 81 - 90 (10)
- 91 or older (11)
- Prefer not to say (12)

Q3 What is your gender?

- Male (1)
- Female (2)
- Prefer to self-describe (3) __________________________________________________
- Prefer not to say (4)

Q12 How long have you been caring for a stroke survivor?

- less than 6 months (1)
- Between 6 months and 1 year (2)
- Between 1 and 5 years (3)
- Between 5 and 10 years (4)
- Over 10 years (5)
- Prefer not to say (6)

Q15 What is your relationship to the stroke survivor?

- Spouse (1)
- Parent (2)
- Sibling (3)
- Other family member (4)
- Friend (5)
- Other (6) __________________________________________________
- Prefer not to say (7)

Q16 Do you live with the stroke survivor?

- No (3)
- Yes (4)
- Prefer not to say (5)

Q17 Are you currently employed?

- No (3)
- Yes (4)
- Prefer not to say (5)

Q18 Overall, how much strain does caring for this person place on you?

- None at all (1)
- A little (2)
- A moderate amount (3)
- A lot (4)
- A great deal (5)
- Prefer not to say (6)

Q13 Do you have access to a car?

- Yes (1)
- No (2)
- Prefer not to say (3)

Q14 How easy or difficult is it for your household to make ends meet financially each month?

- Extremely difficult (1)
- Somewhat difficult (2)
- Somewhat easy (3)
- Extremely easy (4)
- Prefer not to say (5)

Q26 What region do you currently live in?

- North East England (1)
- North West England (2)
- Yorkshire and the Humber (3)
- East Midlands (4)
- West Midlands (5)
- East of England (6)
- London (7)
- South East England (8)
- South West England (9)
- Scotland (10)
- Wales (11)
- Northern Ireland (12)
- Prefer not to say (13)

End of Block: Carer demographics

Start of Block: Clinician demographics

Q44 What is your age?

- 18 - 30 (4)
- 31 - 40 (5)
- 41 - 50 (6)
- 51 - 60 (7)
- 61 - 70 (8)
- 71 - 80 (9)
- 81 - 90 (10)
- 91 or older (11)
- Prefer not to say (12)

Q20 What is your gender?

- Male (1)
- Female (2)
- Prefer to self-describe (3) __________________________________________________
- Prefer not to say (4)

Q21 What is your profession?

- Doctor/ physician (1)
- Physiotherapist (2)
- Occupational therapist (3)
- Speech and language therapist (4)
- Therapy assistant (5)
- Pharmacist (6)
- Nurse (7)
- Dietitian (8)
- Radiographer (9)
- Clinical psychologist (10)
- Physician associate (11)
- Health care assistant (12)
- Other (13) __________________________________________________
- Prefer not to say (14)

Q22 How many years of general clinical experience do you have?

- Less than 1 (1)
- Between 1 and 5 (2)
- Between 6 and 10 (3)
- Between 11 and 15 (4)
- Between 16 and 20 (5)
- More than 20 (6)
- Prefer not to say (7)

Q24 How many years of stroke specific clinical experience do you have?

- Less than 1 (1)
- Between 1 and 5 (2)
- Between 6 and 10 (3)
- Between 11 and 15 (4)
- Between 16 and 20 (5)
- More than 20 (6)
- Prefer not to say (7)

Q27 What region do you currently live in?

- North East England (1)
- North West England (2)
- Yorkshire and the Humber (3)
- East Midlands (4)
- West Midlands (5)
- East of England (6)
- London (7)
- South East England (8)
- South West England (9)
- Scotland (10)
- Wales (11)
- Northern Ireland (12)
- Prefer not to say (13)

End of Block: Clinician demographics

Start of Block: Fitness instructor demographics

Q45 What is your age?

- 18 - 30 (4)
- 31 - 40 (5)
- 41 - 50 (6)
- 51 - 60 (7)
- 61 - 70 (8)
- 71 - 80 (9)
- 81 - 90 (10)
- 91 or older (11)
- Prefer not to say (12)

Q29 What is your gender?

- Male (1)
- Female (2)
- Prefer to self-describe (3) __________________________________________________
- Prefer not to say (4)

Q30 How many years of fitness instructing experience do you have?

- Less than 1 (1)
- Between 1 and 5 (2)
- Between 6 and 10 (3)
- Between 11 and 15 (4)
- Between 16 and 20 (5)
- More than 20 (6)
- Prefer not to say (7)

Q31 What is the highest level qualification related to fitness instructing that you have?

- Level 2 (1)
- Level 3 (2)
- Level 3 (3)
- Level 4 (4)
- Above level 4 (5)
- Prefer not to say (6)

Q32 How much experience do you have working with stroke survivors?

- None at all (1)
- Limited (occasional stroke client) (2)
- Moderate (regular stroke clients) (3)
- Extensive (specialist stroke role) (4)
- Prefer not to say (5)

Q33 What region do you currently live in?

- North East England (1)
- North West England (2)
- Yorkshire and the Humber (3)
- East Midlands (4)
- West Midlands (5)
- East of England (6)
- London (7)
- South East England (8)
- South West England (9)
- Scotland (10)
- Wales (11)
- Northern Ireland (12)
- Prefer not to say (13)

End of Block: Fitness instructor demographics

Start of Block: Survey instructions

Q38 Survey instructions	**It is very important that you read the instructions carefully.**  You will be shown 8 hypothetical scenarios about a potential exercise program. In each scenario, you will be asked to choose between exercise program A, or exercise program B. The two exercise programs will differ by location, price, session frequency, level of individualisation, involvement of carers, and chance of making a meaningful recovery. For both scenarios, the program would **start after discharge from hospital**, be **delivered in a group format by a qualified instructor**, and will last **12 weeks**. Each exercise session will last between **30 - 50 minutes**. Exercises may include walking, cycling on a stationary bike, using resistance bands, lifting weights, and balance exercises.  Below is a description of the different characteristics of the exercise program.  **Location**	This is where the exercise sessions will be delivered. When making your choice, consider how comfortable you are with using technology, the burden of transport, and the amount of support you would need to exercise. The options will be:	At home (connected to the exercise instructor via online software).	At a leisure centre.	Hybrid (a combination of sessions at home and at the leisure centre). **Price**	Price is measured in British pounds (£), per month. As the program is 12 weeks long, the overall price will be equal to the monthly price multiplied by three. The options are:	Free.	£20 per month (£60 for the full 12-week program).	£40 per month (£120 for the full 12-week program). **Session frequency**   This is how many exercise sessions you will take part in per week. The options are:	One per week.	Three per week.	Five per week. 	  **Individualisation**	This is how specific the exercises you are given are to your ability and goals. There are two options.	Targeted group exercises with stroke survivors of similar physical ability.	Generic group exercises with stroke survivors of mixed ability. 	  **Carer involvement**	This is what the carer receives as part of the program. There are three options.	Option one is that carers receive specific education sessions for stroke and exercise, as well as an opportunity to socialise with other carers of stroke survivors.	Option two is that carers take part in the exercise sessions with stroke survivors (at no extra cost), to support the stroke survivor they are caring for.	Option three is that carers are not involved with the program, which may provide some respite, while the stroke survivor is exercising. 	  **Chance of meaningful improvement in recovery**	No intervention can guarantee improvements for stroke survivors, but different types and intensities of rehabilitation can result in a better chance of achieving a positive outcome. A meaningful improvement is something that you would notice in your day-to-day life, for example, to walk up the stairs more easily, be able to use the toilet with more independence. It does not matter what the specific improvement is, as long as it is important and meaningful to you. The options are:	20% chance of making a meaningful recovery. If 10 people took part in this program, 2 of those would achieve a meaningful improvement and 8 would not.	50% chance of making a meaningful recovery. If 10 people took part in this program, 5 of those would achieve a meaningful improvement and 5 would not.	80% chance of making a meaningful recovery. If 10 people took part in this program, 8 of those would achieve a meaningful improvement and 2 would not It is important to note that a 20% chance does not mean a 20% improvement. For exampe, it does not mean that you will be able to walk 20% faster after the program, just that the probability of you achieving your meaningful improvement is 20%.

Q39 I have read and understand these instructions

- Yes (1)
- No (2)

End of Block: Survey instructions

Start of Block: DCE block 1

Q46	.choice-table {	border-collapse: collapse; width: 100%; max-width: 800px; margin-bottom: 1em; font-family: sans-serif; }	.choice-table th, .choice-table td {	border: 1px solid #ccc; padding: 8px; vertical-align: top; text-align: left; }	.choice-table th {	background-color: #f2f2f2; text-align: center; }	**Which of the following exercise programs would you prefer?:** Attribute	Program A	Program B	Location	${lm://Field/1}	${lm://Field/7}	Price	${lm://Field/2}	${lm://Field/8}	Session frequency	${lm://Field/3}	${lm://Field/9}	Individualisation	${lm://Field/4}	${lm://Field/10}	Carer involvement	${lm://Field/5}	${lm://Field/11}	Recovery	${lm://Field/6}	${lm://Field/12}	Click here to view the survey instructions (opens in a new tab)

- Program A (4)
- Program B (5)
- Neither (6)

Display this question:

If Loop current: Which of the following exercise programs would you prefer?: Attribute Program A Program B Locatio... = Neither

Q40 What was your reason for selecting neither?

- I did not like either of the options (1)
- I do not want to do any exercise (2)
- I did not understand the question (3)
- It was too difficult to choose between the scenarios (4)
- I got tired/ bored of answering the questions (5)
- Other (please state) (6) __________________________________________________

End of Block: DCE block 1

Start of Block: DCE block 2

Q39	.choice-table {	border-collapse: collapse; width: 100%; max-width: 800px; margin-bottom: 1em; font-family: sans-serif; }	.choice-table th, .choice-table td {	border: 1px solid #ccc; padding: 8px; vertical-align: top; text-align: left; }	.choice-table th {	background-color: #f2f2f2; text-align: center; }	**Which of the following exercise programs would you prefer?:** Attribute	Program A	Program B	Location	${lm://Field/1}	${lm://Field/7}	Price	${lm://Field/2}	${lm://Field/8}	Session frequency	${lm://Field/3}	${lm://Field/9}	Individualisation	${lm://Field/4}	${lm://Field/10}	Carer involvement	${lm://Field/5}	${lm://Field/11}	Recovery	${lm://Field/6}	${lm://Field/12}	Click here to view the survey instructions (opens in a new tab)

- Program A (1)
- Program B (2)
- Neither (3)

Display this question:

If Loop current: Which of the following exercise programs would you prefer?: Attribute Program A Program B Locatio... = Neither

Q42 What was your reason for selecting neither?

- I did not like either of the options (1)
- I do not want to do any exercise (2)
- I did not understand the question (3)
- It was too difficult to choose between the scenarios (4)
- I got tired/ bored of answering the questions (5)
- Other (please state) (6) __________________________________________________

End of Block: DCE block 2

Start of Block: Future research

Q40 Are you happy to be contacted about future research?

- No (1)
- Yes (2)

Skip To: End of Survey If Are you happy to be contacted about future research? = No

Q41 What is your email address?

________________________________________________________________

End of Block: Future research
